# Automated Phenotypic Characterization in Rare Hematologic Malignancies Using a Large Language Model–Based Framework

**DOI:** 10.64898/2026.06.26.26356633

**Authors:** Muhammad Ali Khan, Umair Ayub, Salman Ayub Jajja, Muhammad Umair Anjum, Kainat Warraich, Prateek Jain, Japneet Kaur Oberoi, Mousab Al Abbas, Muhammad Hussnain Sadiq, Muhammad Uzair Sarfraz, Ziyi Huang, Irbaz Bin Riaz, Jeanne M. Palmer

## Abstract

**Background:** Diagnosis and risk stratification in rare hematologic malignancies such as myeloproliferative neoplasms (MPNs) – polycythemia vera (PV), essential thrombocythemia (ET), and myelofibrosis (MF) – require expert review of longitudinal, heterogeneous clinical records. This process is cognitively demanding, inconsistently applied, and difficult to scale beyond tertiary centers. No automated phenotyping workflow currently exists for hematologic malignancies.

**Methods:** A HIPAA-compliant large language model (LLM) framework for phenotyping MPN was developed to integrate (i) rule-based retrieval of bone marrow biopsy reports, clinical notes, and structured laboratory results from the electronic health record (EHR); (ii) zero-shot extraction of diagnostic and prognostic variables from unstructured text using GPT-4 Turbo; (iii) a clinician-informed source-prioritization algorithm to reconcile conflicting multi-source data; (iv) WHO/ICC-criteria–based diagnostic classification; and (v) NCCN-based risk stratification using the conventional risk model for PV, IPSET-thrombosis for ET, and DIPSS, DIPSS-plus, and MIPSS70/MIPSS70+ v2 for MF. Patients were identified via MPN-related ICD-9/10 codes; cases met 2017 WHO criteria or had a hematologist-documented diagnosis, and controls did not. The cohort was split into a prompt-development set (n = 60) and a held-out test set (n = 450; 75 cases and 75 controls per disease). Ground truth was established by independent dual-clinician chart review with consensus adjudication. LLM performance was evaluated against the ground truth: variable-level extraction using accuracy, F1 score, and Cohen’s kappa; patient-level diagnostic classification using sensitivity, specificity, and Cohen’s kappa; and prognostic risk stratification (among confirmed cases) using accuracy, weighted F1 score, and quadratic-weighted Cohen’s kappa. Wilson 95% confidence intervals (CIs) were used for proportions and bootstrap 95% CIs with 500 resamples for F1 scores.

**Results:** The held-out test set included 450 patients (PV: 150; ET: 150; MF: 150) with pathology reports and structured laboratory results, and 172 patients (PV: 52; ET: 55; MF: 65) with clinical notes. From pathology reports, overall variable extraction accuracy and F1 score were 99% (95% CI, 98–100) and 1.00 (0.99–1.00) for PV, 100% (99–100) and 0.99 (0.96–1.00) for ET, and 100% (99–100) and 0.99 (0.97–1.00) for MF. From clinical notes, overall accuracy and F1 score were 96% (91–100) and 0.94 (0.85–1.00) for PV, 100% (100–100) and 1.00 (1.00–1.00) for ET, and 100% (99–100) and 0.98 (0.95–1.00) for MF. Diagnostic sensitivity was 100% (95% CI, 95.1–100.0) for PV, ET, and MF; specificity was 98.7% (92.8–99.8) for PV and 100% (95.1–100.0) for both ET and MF, with Cohen’s kappa of 0.99 for PV and 1.00 for ET and MF. Risk stratification accuracy was 100% with weighted F1 score of 1.00 and quadratic-weighted Cohen’s kappa of 1.00 across all three diseases. A pre-specified source-ablation analysis showed that pathology reports alone were sufficient for diagnosis (sensitivity 98.7% for PV, 100% for ET, 96.0% for MF; specificity 100% across all three subtypes) but inadequate for prognostication (accuracy 69.3% for PV, 93.3% for ET, 77.3% for MF). Adding clinical notes to pathology reports recovered full prognostic accuracy of 100% across all three diseases.

**Conclusions:** This first-in-class automated framework achieved expert-level performance for MPN diagnosis and risk stratification from real-world EHR data, establishing a foundation for scalable, standardized phenotyping in rare hematologic malignancies. Prospective, multi-site validation is warranted before clinical deployment.

## Introduction

Hematologic malignancies pose a fundamental challenge for automated phenotypic characterization. Unlike many solid tumors, in which diagnosis and prognostication are anchored to a discrete biopsy, the information needed for these tasks in hematologic malignancies accumulates gradually and is distributed across structured laboratory results, molecular testing reports, pathology, imaging studies, and longitudinal clinical documentation generated across repeated encounters. The nonspecificity of early findings adds further complexity, particularly for rare hematologic malignancies that often sit low on a differential diagnosis list. Manual chart review remains the prevailing approach to this synthesis, despite being labor-intensive, cognitively demanding, and difficult to scale.^1,2^ Despite rapid advances in clinical artificial intelligence,^3,4^ no automated phenotyping workflow currently exists for hematologic malignancies.

Myeloproliferative neoplasms (MPNs) i.e., polycythemia vera (PV), essential thrombocythemia (ET), and myelofibrosis (MF), exemplify this broader challenge. MPNs are clonal hematologic malignancies that collectively affect approximately 2 to 3 individuals per 100,000 person-years and are associated with substantial morbidity, excess mortality, thrombotic events, progressive marrow fibrosis, and leukemic transformation.^5–10^ Their recognition is often delayed because early manifestations like incidental thrombocytosis, unexplained erythrocytosis, fatigue, pruritus, splenomegaly, and constitutional symptoms overlap with benign hematologic abnormalities and common comorbidities.^7,11^ Diagnosis requires synthesis of complete blood counts, serum erythropoietin and lactate dehydrogenase concentrations, bone marrow morphology and reticulin fibrosis grade, JAK2/CALR/MPL mutation status, splenomegaly assessment, and exclusion of reactive cytoses according to World Health Organization (WHO) and International Consensus Classification (ICC) criteria.^12,13^

Following diagnosis, treatment decisions depend on subtype-specific risk stratification. The conventional PV model requires age and prior thrombotic history; IPSET-thrombosis for ET requires age, thrombotic history, cardiovascular risk factors, and JAK2 mutation status; and MF models i.e., DIPSS, DIPSS-plus, MIPSS70, and MIPSS70+ v2 require age, constitutional symptoms, hemoglobin, leukocyte count, circulating blast percentage, platelet count, transfusion dependence, cytogenetics, and high-molecular-risk mutations (ASXL1, SRSF2, EZH2, IDH1/2, U2AF1).^14–19^ These variables typically require reconciliation across pathology reports, molecular results, clinical notes, and longitudinal laboratory data. Incomplete phenotyping can therefore delay diagnosis, distort risk assignment, and forgo evidence-based interventions, consequences that are particularly common in community oncology settings, where most patients with MPN receive care and where limited subspecialty exposure and variable documentation practices further constrain systematic recognition.^20,21^ Existing natural language processing and large language model (LLM) approaches in oncology have shown remarkable data abstraction capabilities across various document types,^22–24^ however, these models are largely focused on solid tumors, where diagnostic anchors and staging variables are concentrated in pathology and radiology reports or structured registries,^25–27^ and do not address the longitudinal, multi-source reasoning required for hematologic phenotyping.

To address this gap, we developed and evaluated a HIPAA-compliant, LLM-based framework for automated phenotypic characterization of MPNs. The pipeline integrates rule-based retrieval of relevant electronic health record documents, zero-shot extraction of diagnostic and prognostic variables from unstructured clinical text using GPT-4 Turbo, clinician-informed source prioritization to reconcile conflicting data, diagnostic classification according to WHO/ICC criteria, and subtype-specific risk stratification using established prognostic models. MPNs were chosen as a proof-of-concept because they require the same core capabilities needed for scalable phenotyping across rare hematologic malignancies: longitudinal data integration, multi-source evidence reconciliation, subtype classification, and prognostic variable extraction. We hypothesized that this framework would enable accurate, reproducible, and scalable MPN phenotyping while providing a template adaptable to other rare hematologic malignancies with complex diagnostic and prognostic criteria.

## Methods

### Study Design

We conducted a retrospective study to develop and evaluate an end-to-end LLM framework for automated phenotypic characterization of MPNs, encompassing diagnostic classification and prognostic risk stratification. Patients were drawn from across all Mayo Clinic sites.

### Data Source and Study Population

Patients were identified from the institutional EHR using International Classification of Diseases, Ninth and Tenth Revision (ICD-9 and ICD-10) codes corresponding to PV, ET, and MF. Cases were defined as patients who fulfilled the 2017 WHO diagnostic criteria for PV, ET, or MF based on clinical, laboratory, and pathological findings, or who had a definitive hematologist-documented diagnosis in the clinical record. Controls were defined as patients with MPN-related ICD codes who did not meet WHO diagnostic criteria and did not have a hematologist-confirmed diagnosis of MPN. Patients without a documented bone marrow biopsy were excluded.

A random sample of 510 patients was identified, comprising 85 cases each of PV, ET, and MF and 255 matched controls (85 per disease). Sixty patients were allocated to a prompt-development set; the remaining 450 (75 cases and 75 controls per disease) were used as a locked, held-out test set used exclusively for performance evaluation.

### Ground Truth Development

Two clinicians independently reviewed clinical notes and pathology reports for each patient and abstracted demographic variables, clinical features (including symptoms and thrombotic history), laboratory values (including complete blood counts, serum erythropoietin, and lactate dehydrogenase), genetic information (driver and additional mutations and karyotype), bone marrow morphology findings, diagnostic documentation, and prognostic-score variables. The appropriate prognostic model was selected based on confirmed diagnosis and availability of required variables: the conventional risk model for PV; IPSET-thrombosis for ET; and DIPSS, DIPSS-plus, MIPSS70, or MIPSS70+ v2 for MF. Disagreements between reviewers were resolved by a third clinician through consensus review.

### Automated Workflow

The automated framework consisted of six sequential components, summarized schematically in **Figure 1**.

***1. Document retrieval.*** A rule-based algorithm retrieved key clinical documents from the EHR for each patient, including basic demographic information, the first bone marrow biopsy pathology report, the first clinical note generated after that biopsy, and the most recent set of structured laboratory results within a 90-day window before the biopsy date.
***2. LLM-based data extraction.*** A HIPAA-compliant deployment of GPT-4 Turbo, accessed through an institutional Apigee-proxied Azure OpenAI endpoint, was used to extract demographic, clinical, laboratory, genetic, and pathological variables from unstructured clinical notes and pathology reports. Structured prompts were developed using the prompt-development set and then frozen prior to any evaluation on the held-out test set. Model inference was performed under deterministic decoding settings.
***3. Data post-processing.*** Regular-expression-based scripts converted free-text LLM outputs into structured tabular variables and harmonized categorical and numeric formatting across documents.
***4. Source prioritization.*** A clinician-informed hierarchical algorithm was applied to reconcile values across data sources when multiple values were available for the same variable. The hierarchy gave precedence to structured laboratory results for numeric variables, to pathology reports for morphology and pathology variables, and to clinical notes for symptom and history variables, with explicit fall-through rules and missingness handling. Decision trees implementing the source-prioritization logic for categorical (morphology, karyotype, and mutation) variables and for numeric laboratory variables are provided in Supplementary Figures 1 and 2, respectively.
***5. Diagnostic classification.*** A rule-based classifier implementing WHO and ICC criteria was applied to distinguish MPN cases from controls and assign disease subtype. Hematologist-documented diagnoses were incorporated when available and treated as confirmatory.
***6. Risk stratification.*** An NCCN-based algorithm selected the appropriate prognostic model and computed risk categories using available clinical variables: low/high risk for PV (conventional risk model); very-low, low, intermediate, or high risk for ET (IPSET-thrombosis); and lower (MF-1) versus higher (MF-2) risk for MF, where the choice between DIPSS/DIPSS-plus and MIPSS70/MIPSS70+ v2 depended on the availability of cytogenetic and high-molecular-risk mutation data.

**Figure 1.**
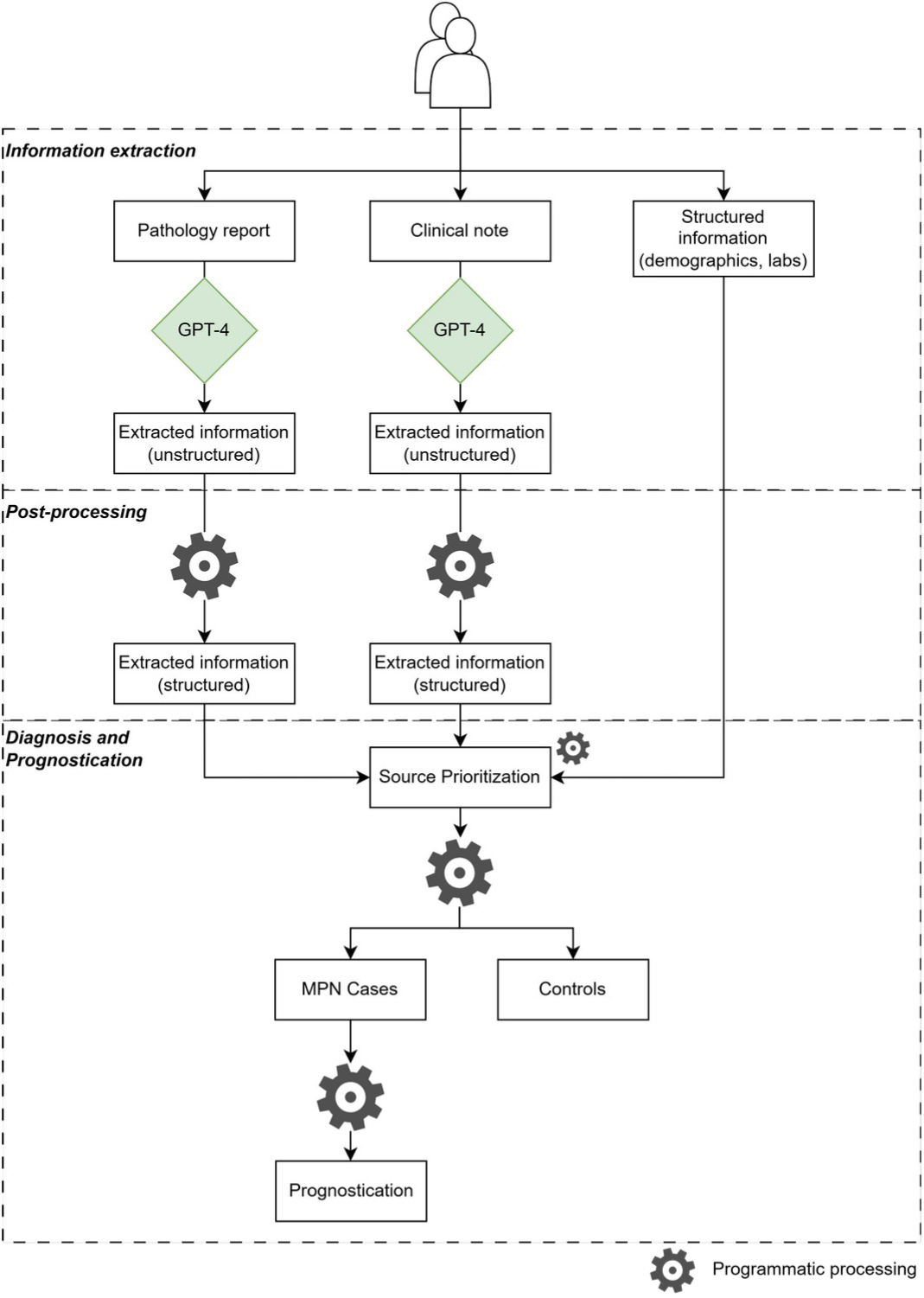
Schematic overview of the automated MPN phenotyping pipeline. LLM-based framework for MPN phenotyping. The pipeline comprises three stages, indicated by dashed dividers. **Information extraction:** rule-based retrieval supplies pathology reports, clinical notes, and structured EHR information (demographics and laboratory results); GPT-4 Turbo extracts diagnostic and prognostic variables from each unstructured document type into a free-text representation. **Post-processing:** regular-expression-based scripts convert the free-text outputs into structured tabular variables harmonized across documents. **Diagnosis and prognostication:** a clinician-informed source-prioritization algorithm reconciles values across data sources; rule-based logic implementing WHO and ICC criteria classifies patients as MPN cases or controls; and confirmed cases are passed to a subtype-specific prognostic module that selects between the conventional risk model (PV), IPSET-thrombosis (ET), and DIPSS, DIPSS-plus, and MIPSS70/MIPSS70+ v2 (MF). Decision trees for the source-prioritization step are provided in Supplementary Figures 1 (categorical variables) and 2 (numeric laboratory variables). **Abbreviations:** DIPSS, Dynamic International Prognostic Scoring System; EHR, electronic health record; ET, essential thrombocythemia; ICC, International Consensus Classification; IPSET, International Prognostic Score for Thrombosis in Essential Thrombocythemia; LLM, large language model; MF, myelofibrosis; MIPSS70, Mutation-Enhanced International Prognostic Score System; MPN, myeloproliferative neoplasm; PV, polycythemia vera; WHO, World Health Organization.

**Figure 2.**
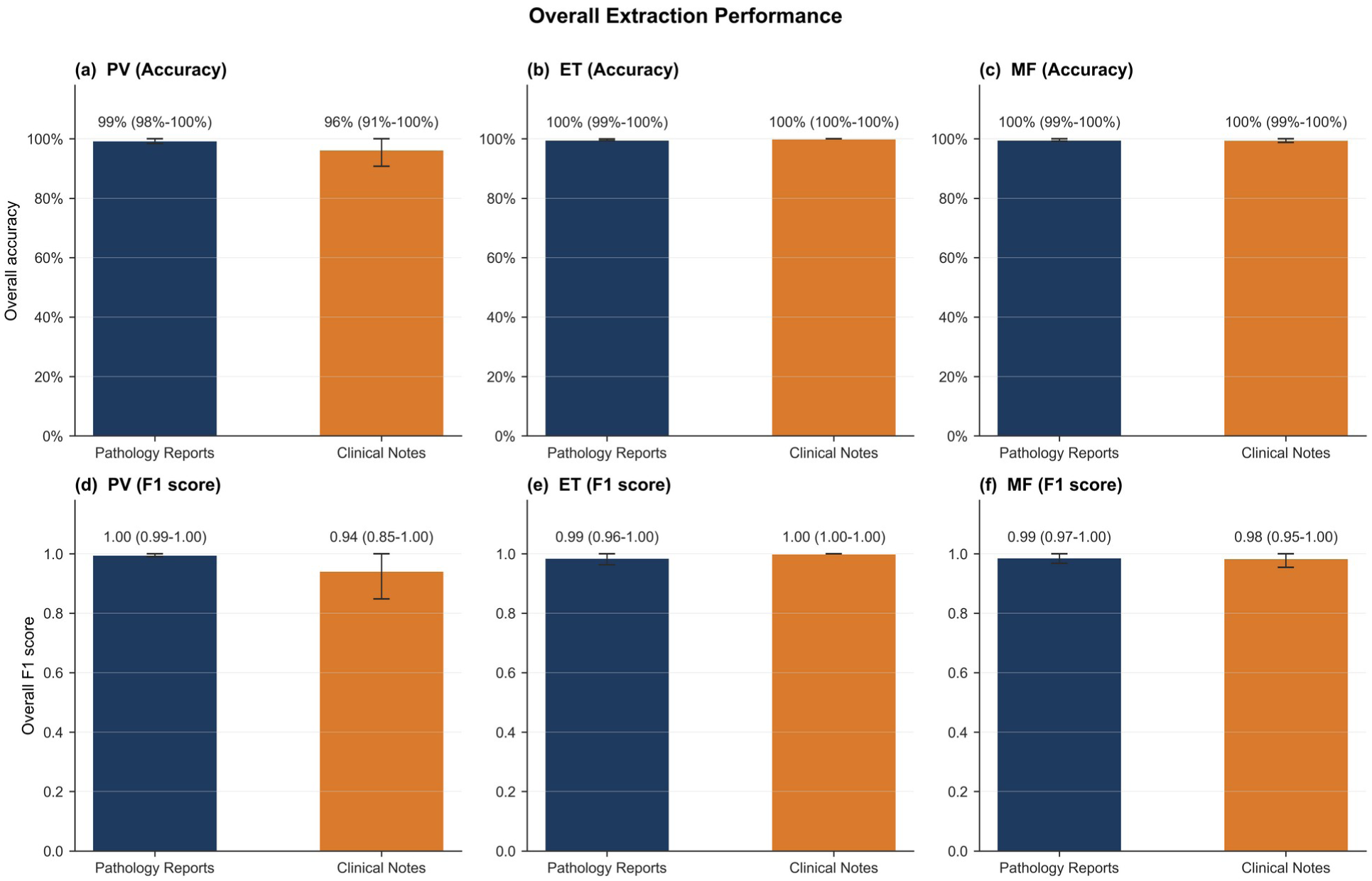
Overall LLM extraction performance versus manual abstraction by disease and document type. Bar plots show overall accuracy (top row, panels a–c) and overall F1 score (bottom row, panels d–f) for LLM-derived versus manually abstracted variables, separately for pathology reports (navy) and clinical notes (orange), in PV (a, d), ET (b, e), and MF (c, f). Numbers above each bar give the point estimate and 95% confidence interval. Pathology-report extraction accuracy was 99% for PV and 100% for ET and MF; clinical-note extraction accuracy was 96% for PV and 100% for ET and MF. **Abbreviations:** ET, essential thrombocythemia; F1, F1 score; LLM, large language model; MF, myelofibrosis; PV, polycythemia vera.

### Statistical Analysis

Performance of automated extraction, diagnostic classification, and risk stratification was evaluated against the manually curated ground-truth dataset. For variable extraction, paired comparisons were made between LLM-derived values and manually abstracted values per document and per variable; accuracy, F1 score, and Cohen’s kappa were computed. For diagnostic classification, sensitivity, specificity, and Cohen’s kappa were computed at the patient level; positive and negative predictive values were not used as primary metrics because the case-to-control sampling ratio was fixed at 1:1 and does not reflect the true population prevalence of MPN. For risk stratification, accuracy, weighted F1 score, and quadratic-weighted Cohen’s kappa were computed on confirmed cases. Wilson 95% confidence intervals (CIs) were used for proportions, and bootstrap 95% CIs with 500 resamples were used for F1 score. Because of the limited interpretability of small-denominator CIs at the per-variable level, 95% CIs are reported only for aggregate disease-level extraction performance (Figure 2) and for patient-level diagnostic, prognostic, and source-ablation metrics (Supplementary Tables 2–4); per-variable extraction values are reported as point estimates accompanied by paired-observation denominators (Supplementary Table 1). A pre-specified source-ablation analysis evaluated four progressively richer source configurations, pathology only, pathology + clinical notes, pathology + structured labs, and all sources, to characterize the marginal contribution of each data modality. All analyses were performed in Python.

### Ethics

The study was approved by the institutional review board (IRB # 21-002040) with a waiver of informed consent. All data were handled in accordance with Health Insurance Portability and Accountability Act (HIPAA) regulations, and no patient-identifiable information was transferred outside the institution’s secure computing environment.

## Results

### Study Population

A total of 510 patients with MPN-related ICD codes and a documented bone marrow biopsy were included. Sixty patients were allocated to the prompt-development set, and 450 patients were held out for evaluation. The held-out test set comprised 75 cases and 75 controls per disease (n = 150 per disease; n = 450 in total). Baseline characteristics are summarized in **Table 1**. Pathology reports and structured laboratory results were available for all 450 patients (PV: 150; ET: 150; MF: 150); clinical notes were available for 172 patients (PV: 52; ET: 55; MF: 65). Median pathology-report length ranged from approximately 1,260 to 1,688 tokens across groups, and median clinical-note length ranged from approximately 1,064 to 3,036 tokens, computed using the OpenAI cl100k_base tokenizer.

**Table 1.** Baseline characteristics of the held-out test set (n = 450).

Cohort characteristics for the locked, held-out evaluation set comprising 75 cases and 75 controls per disease.
|  | Polycythemia vera |  | Essential thrombocythemia |  | Myelofibrosis |  |
| --- | --- | --- | --- | --- | --- | --- |
| Variable | Cases | Controls | Cases | Controls | Cases | Controls |
| N (patients) | 75 | 75 | 75 | 75 | 75 | 75 |
| Age, median (IQR), years | 63.9 (54.9–72.1) | 64.8 (52.2–74.1) | 59.1 (48.9–69.1) | 69.0 (62.5–76.8) | 69.3 (61.5–76.0) | 69.1 (57.4–75.8) |
| Male sex, n (%) | 43 (57.3) | 42 (56.0) | 20 (27.0) | 39 (52.0) | 37 (49.3) | 47 (62.7) |
| Pathology reports per patient, median (IQR) | 1 (1–1) | 1 (1–1) | 1 (1–1) | 1 (1–1) | 1 (1–1) | 1 (1–1) |
| Clinical notes per patient, median (IQR) | 0 (0–1) | 0 (0–0) | 1 (0–1) | 0 (0–0) | 1 (0–1) | 0 (0–0) |
| Structured lab sets per patient, median (IQR) | 1 (1–1) | 1 (1–1) | 1 (1–1) | 1 (1–1) | 1 (1–1) | 1 (1–1) |
| Pathology report tokens, median (IQR) | 1490 (1377–1630) | 1260 (1010–1516) | 1513 (1400–1676) | 1374 (1041–1636) | 1688 (1478–1892) | 1414 (1110–1689) |
| Clinical note tokens, median (IQR) | 1511 (810–2105) | 1172 (602–2198) | 1064 (527–1816) | 1337 (934–2334) | 1391 (1106–2046) | 3036 (1523–4516) |

### Variable Extraction Performance

For pathology reports, overall accuracy and F1 score were 99% (95% CI, 98–100) and 1.00 (95% CI, 0.99–1.00) for PV, 100% (99–100) and 0.99 (0.96–1.00) for ET, and 100% (99–100) and 0.99 (0.97–1.00) for MF (**Figure 2**). For clinical notes, overall accuracy and F1 score were 96% (91–100) and 0.94 (0.85–1.00) for PV, 100% (100–100) and 1.00 (1.00–1.00) for ET, and 100% (99–100) and 0.98 (0.95–1.00) for MF.

Variable-level performance by disease is shown in **Figure 3A–C** and detailed in Supplementary Table 1. For categorical mutations (JAK2, CALR, MPL, ASXL1, EZH2, TET2, IDH1/2, SRSF2, SF3B1), accuracy ranged from 99.3% to 100% from pathology reports and clinical notes. For categorical morphology variables (hypercellularity, megakaryocyte proliferation, megakaryocyte atypia, reticulin fibrosis grade, trilineage hematopoiesis), accuracy ranged from 97.9% to 100.0% across both document types. For numeric variables (hemoglobin, hematocrit, platelet count, white blood cell count, peripheral blood blasts, percent cellularity, erythropoietin), accuracy was 99.3% to 100.0% in pathology reports and 81.0% to 100.0% in clinical notes. Hematologist-defined disease labels in PV had accuracy of 99.3% and F1 score of 0.99 in pathology reports and 94.2% and 0.96 in clinical notes. The lowest single-variable performance occurred for hematocrit extracted from PV clinical notes, with accuracy of 81.0% and F1 score of 0.68 (n = 21). Concordance between structured laboratory values and pathology-report-extracted numeric values was as follows: in PV, Pearson r = 1.00 for hemoglobin and 1.00 for hematocrit (Supplementary Figure 3); in ET, r = 0.92 for platelet count, 0.99 for hemoglobin, and 1.00 for white blood cell count (Supplementary Figure 4); in MF, r = 0.99 for hemoglobin, 1.00 for white blood cell count, and 0.97 for peripheral blood blasts (Supplementary Figure 5).

**Figure 3.**
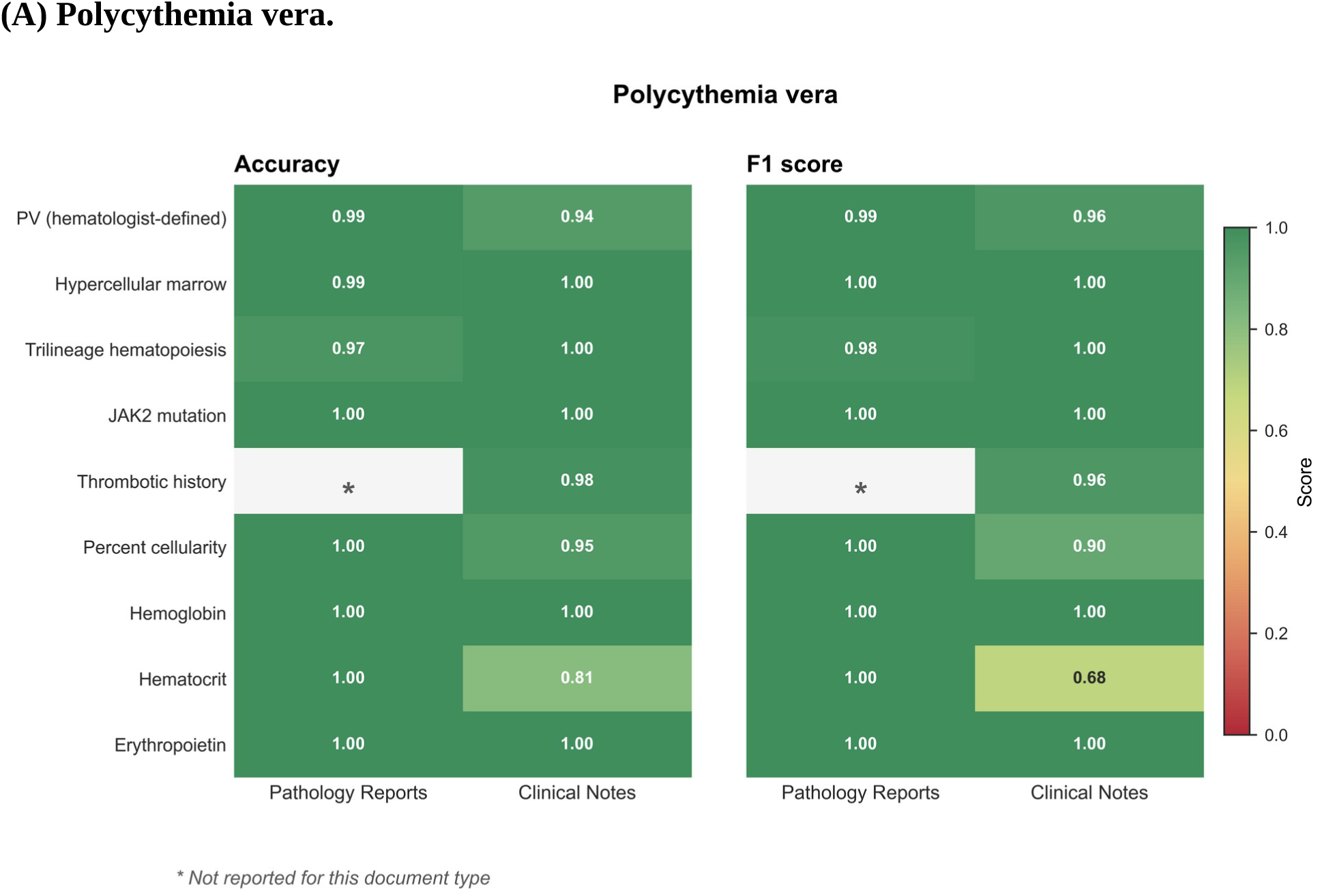
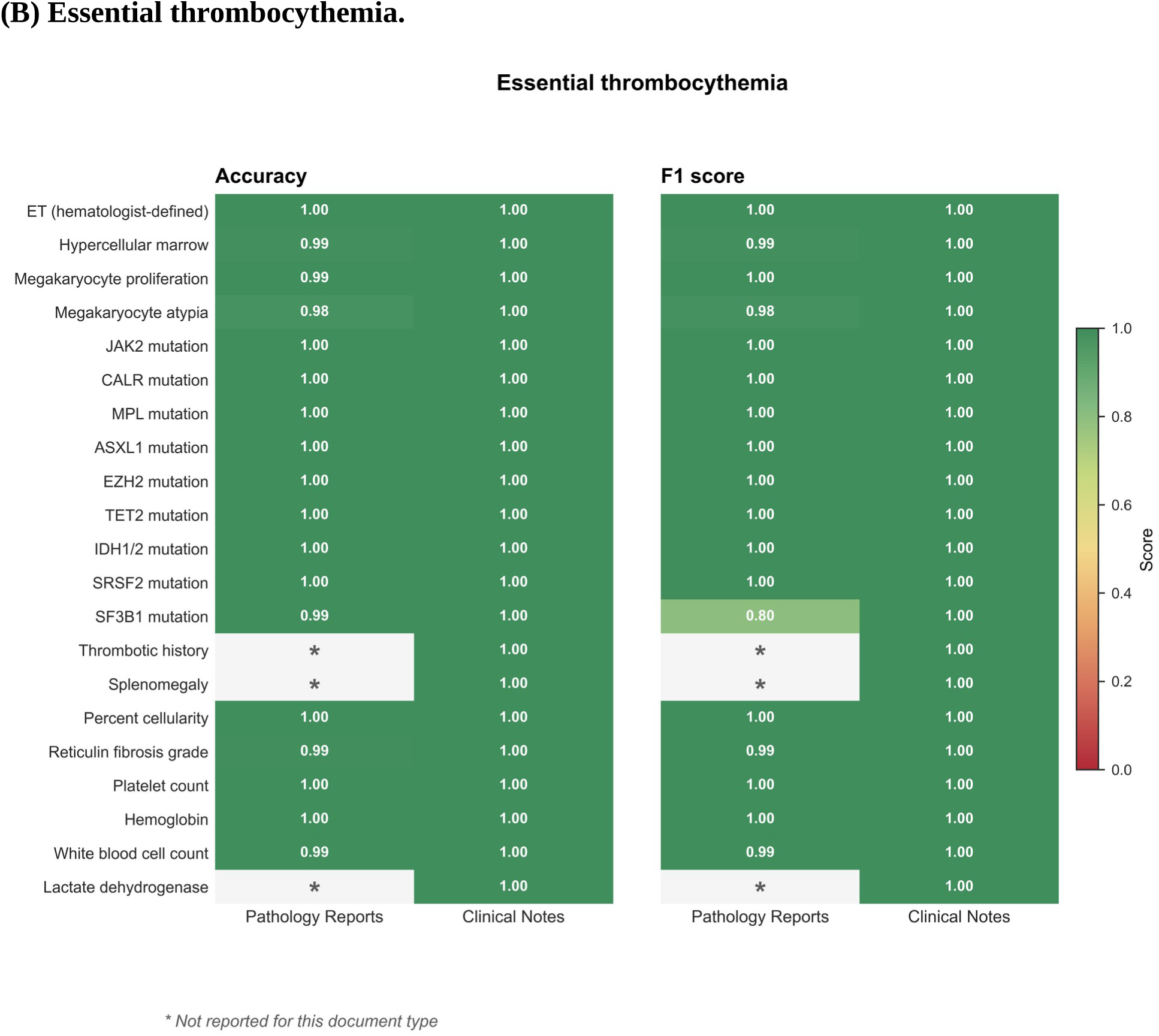
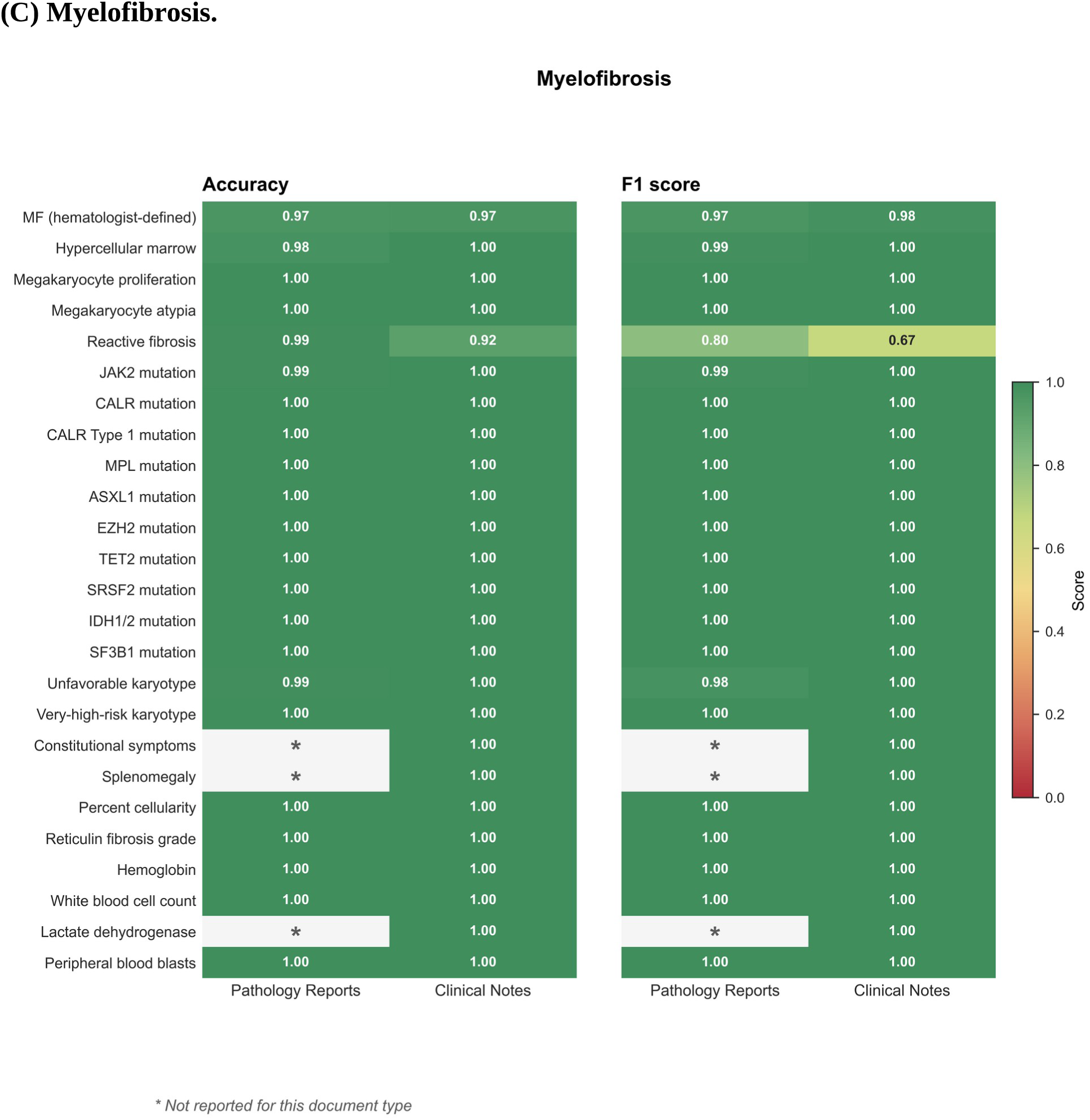
Variable-level LLM extraction accuracy and F1 score by disease and document type. Heatmaps display per-variable accuracy (left) and F1 score (right) for LLM-extracted versus manually abstracted values, separately for pathology reports and clinical notes. Cells are coloured on a continuous gradient from red (poor) to green (good), with values printed inside each cell. Asterisks (*) denote variables that were not reported in the corresponding document type. (A) PV. (B) ET. (C) MF. The single low-F1 cell for ET pathology-report SF3B1 (F1 = 0.80) and for MF reactive-fibrosis (F1 = 0.80 from pathology, 0.67 from notes) reflects very low positive prevalence; the single low-F1 cell in PV clinical notes (hematocrit; F1 = 0.68) reflects a small denominator (n = 21) due to inconsistent free-text reporting. Per-variable point estimates and Cohen’s kappa are provided in Supplementary Table 1. **Abbreviations:** ET, essential thrombocythemia; F1, F1 score; MF, myelofibrosis; PV, polycythemia vera.

### Diagnostic Classification Performance

Patient-level diagnostic performance is summarized in **Figure 4** and Supplementary Table 2. The framework achieved 100% sensitivity (75/75; 95% CI, 95.1–100.0) for PV, ET, and MF.

**Figure 4.**
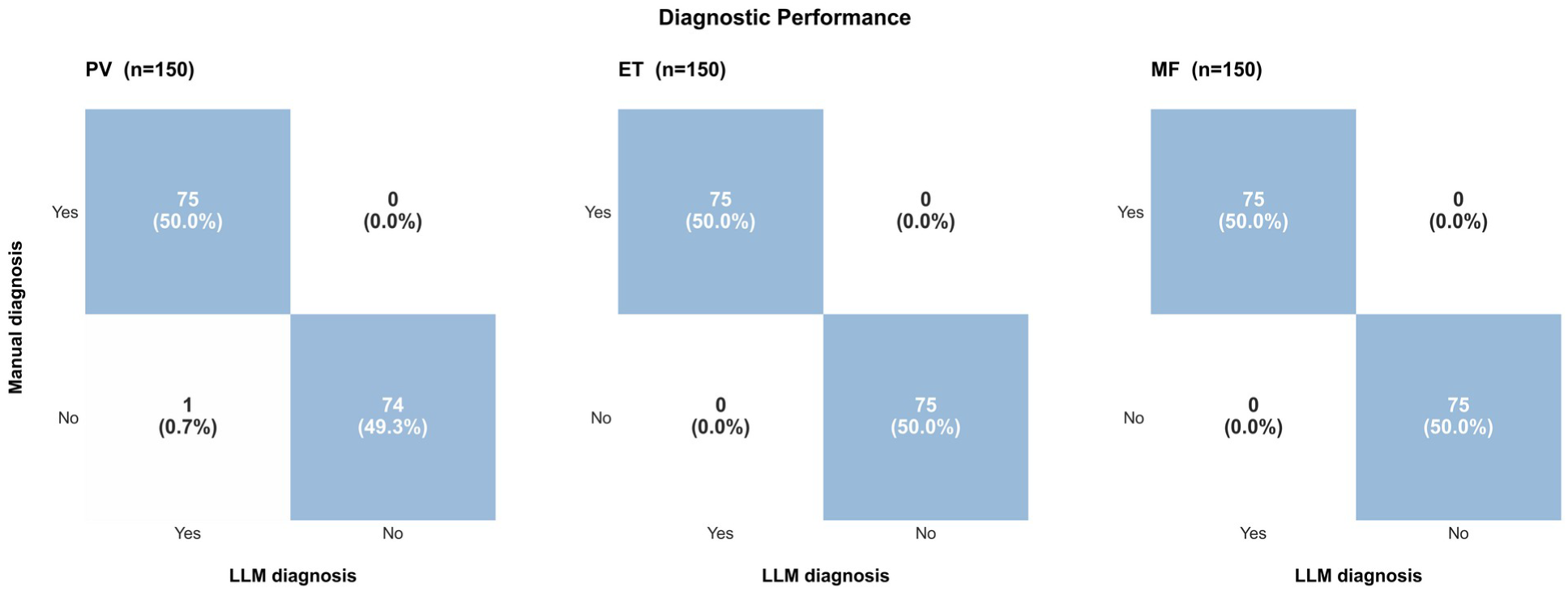
LLM versus manual diagnosis: confusion matrices. Confusion matrices comparing LLM-derived diagnosis (columns) with the manually adjudicated reference standard (rows), shown for PV (left), ET (middle), and MF (right) in the held-out test set (n = 150 per disease, 75 cases and 75 controls). Cell labels show counts and percentages of total. The framework achieved 100% sensitivity (95% CI, 95.1–100.0) for all three diseases. Specificity was 98.7% (95% CI, 92.8–99.8) for PV, corresponding to one false-positive control among 75, and 100% (95.1–100.0) for ET and MF. **Abbreviations:** ET, essential thrombocythemia; LLM, large language model; MF, myelofibrosis; PV, polycythemia vera.

Specificity was 98.7% (74/75; 95% CI, 92.8–99.8) for PV and 100% (75/75; 95.1–100.0) for both ET and MF. Cohen’s kappa was 0.99 for PV and 1.00 for ET and MF.

### Risk Stratification Performance

Among confirmed cases (n = 75 per disease), risk stratification was concordant with manually adjudicated risk in 100% of patients (75/75; 95% CI, 95.1–100.0) for all three subtypes, with weighted F1 score of 1.00 (95% CI, 1.00–1.00) and quadratic-weighted Cohen’s kappa of 1.00 (**Figure 5**; Supplementary Table 3). For PV, all 9 patients adjudicated as low-risk and all 66 as high-risk were correctly categorized. For ET, all 20 patients adjudicated as very low risk, 14 as low risk, 11 as intermediate risk, and 29 as high risk were correctly categorized. For MF, all 15 patients in the lower-risk (MF-1) tier and all 60 in the higher-risk (MF-2) tier were correctly categorized; the framework selected among DIPSS, DIPSS-plus, and MIPSS70/MIPSS70+ v2 based on the availability of cytogenetic and high-molecular-risk mutation data.

**Figure 5.**
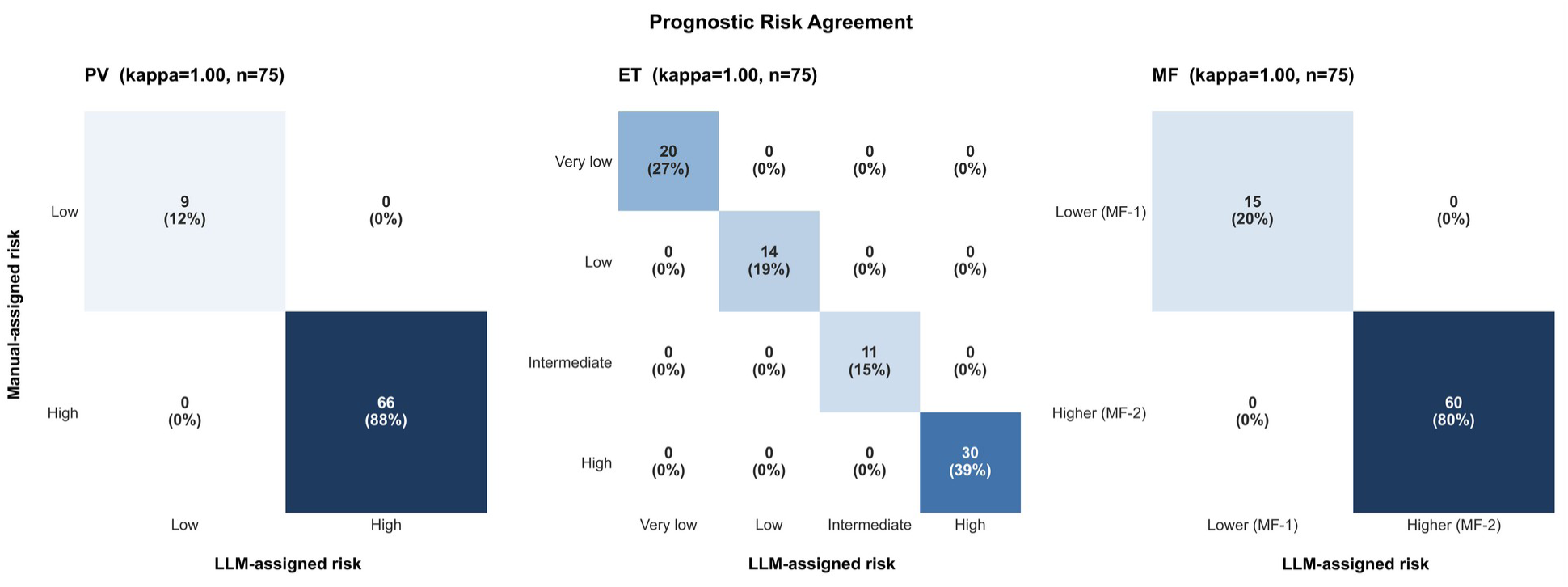
Prognostic risk agreement between LLM-assigned and manually assigned risk categories. Confusion matrices comparing LLM-assigned prognostic risk (columns) with the manually adjudicated risk category (rows), shown for confirmed cases of PV (left; 2-tier conventional risk model), ET (middle; 4-tier IPSET-thrombosis), and MF (right; 2-tier MIPSS-based stratification with selection between DIPSS, DIPSS-plus, and MIPSS70/MIPSS70+ v2 according to data availability). All three matrices are diagonal; quadratic-weighted Cohen’s kappa was 1.00 for each disease (n = 75 cases per disease). Cell labels show counts and the percentage of confirmed cases per disease. **Abbreviations:** DIPSS, Dynamic International Prognostic Scoring System; ET, essential thrombocythemia; IPSET, International Prognostic Score for Thrombosis in Essential Thrombocythemia; LLM, large language model; MF, myelofibrosis; MIPSS70, Mutation-Enhanced International Prognostic Score System; MF-1, lower-risk myelofibrosis; MF-2, higher-risk myelofibrosis; PV, polycythemia vera.

### Source Ablation Analysis

A pre-specified source-ablation analysis evaluated each EHR data modality (Supplementary Figures 6–7; Supplementary Table 4). With pathology reports alone, diagnostic sensitivity was 98.7% for PV, 100% for ET, and 96.0% for MF; specificity was at or near 100% for all three subtypes. Prognostic accuracy with pathology reports alone was 69.3% for PV, 93.3% for ET, and 77.3% for MF. Adding structured laboratory data did not change prognostic accuracy for PV (69.3%) or ET (93.3%) and increased it modestly for MF (82.7%). Adding clinical notes to pathology reports increased prognostic accuracy to 100% across all three diseases. The pathology + clinical notes + structured laboratory data configuration yielded the same prognostic accuracy as the pathology + clinical notes configuration.

## Discussion

This study developed and evaluated an end-to-end LLM-based framework for automated phenotypic characterization of MPNs from real-world EHR data. Across the held-out test set of 450 patients, variable extraction achieved overall accuracy of 96% to 100% from pathology reports and clinical notes; the framework correctly classified 100% of cases as PV, ET, or MF (sensitivity 100%; specificity 98.7% for PV and 100% for ET and MF) and assigned the correct prognostic risk category in 75 of 75 cases per disease (weighted F1 score 1.00; quadratic-weighted Cohen’s kappa 1.00). To our knowledge, this is the first automated phenotyping framework demonstrated for a rare hematologic malignancy and shows that the multi-step diagnostic and prognostic reasoning required for MPN classification, spanning laboratory, morphologic, molecular, and narrative data, is amenable to LLM-based automation.

Several observations warrant comment. Extraction accuracy in clinical notes was lower than in pathology reports for PV (96% vs. 99%), driven primarily by hematocrit (81.0%; n = 21) and hematologist-defined disease labels (94.2%; n = 52); these likely reflect inconsistent free-text reporting and greater linguistic variability in narrative documentation. SF3B1 was assessed in ET because myelodysplastic/myeloproliferative neoplasm with ring sideroblasts and thrombocytosis (MDS/MPN-RS-T; historically termed RARS-T), which is characterized by SF3B1 mutation, can present with thrombocytosis that mimics ET; when such patients also carry a JAK2 mutation, they may retain an MPN-like thrombotic risk, so capturing SF3B1 status helps flag this overlap. For SF3B1 in ET pathology reports, only 3 of 150 cases were positive, so a single misclassification yielded 99.3% accuracy but reduced F1 score to 0.80. The source-ablation analysis showed that pathology reports alone were sufficient for diagnosis (sensitivity 96.0%–100%; specificity 100%) but yielded prognostic accuracy of only 69.3%–93.3%; adding clinical notes restored prognostic accuracy to 100%, whereas adding structured laboratory data alone provided minimal incremental gain. This is consistent with prognostic models depending on variables such as thrombotic history, constitutional symptoms, and splenomegaly that are typically captured in narrative documentation rather than discrete EHR fields.

Diagnostic performance compares favorably with previously reported NLP- and LLM-based phenotyping systems in oncology,^28,29^ which have largely focused on solid tumors with discrete diagnostic events and structured pathology reports. In contrast, the present framework integrates structured laboratory data, pathology reports, and longitudinal clinical narratives within a unified workflow that explicitly applies WHO and ICC criteria^12,13^ to reconcile extracted variables and assign disease subtype. The framework also selects among MF prognostic models (DIPSS, DIPSS-plus, MIPSS70/MIPSS70+ v2)^18,19^ based on the availability of cytogenetic and high-molecular-risk mutation data. Within our cohort, prognostic stratification was effectively binary for PV (low vs. high risk) and MF (MF-1 vs. MF-2) and four-class for ET; performance depended on the upstream availability of the constituent variables, and source ablation showed that prognostic accuracy fell to 69%–93% in the absence of clinical notes, with most errors attributable to incomplete documentation rather than extraction failure. This suggests that the principal barrier to automated MPN risk stratification in real-world settings is data availability rather than computational capability, and that deployment will need to address the heterogeneity of how prognostic variables are documented across institutions.

The potential clinical implications of this framework are substantial. Accurate MPN phenotyping requires careful review of longitudinal records and familiarity with disease-specific diagnostic and prognostic criteria. Outside tertiary referral centers, community oncologists often manage diverse malignancies and may have limited exposure to MPN-specific workflows.^20,21^ Automated tools capable of retrieving relevant documents, extracting key variables, and applying standardized diagnostic and prognostic logic could reduce diagnostic delays and improve the completeness of risk stratification in routine practice.

The strengths of this study include a locked, prespecified held-out test set; dual-clinician ground-truth adjudication; deterministic LLM inference with prompts frozen prior to evaluation; reporting of metrics aligned to the structure of each task (extraction, binary diagnostic classification, ordinal prognostic stratification); and a prespecified source-ablation analysis that quantifies the contribution of each EHR data modality. The framework was developed under HIPAA-compliant conditions and required no model fine-tuning, supporting the feasibility of zero-shot deployment in institutions with access to a hosted LLM.

However, the findings of the study should be interpreted in the context of specific limitations. First, the cohort was drawn from a single multi-site health system, and performance across institutions with different EHR systems and documentation conventions is unknown.^30,31^ Second, cases were oversampled relative to the population prevalence of MPNs; positive and negative predictive values are not informative under this sampling design and were not reported. Third, the retrospective design assumes record completeness at the time of evaluation, whereas real-time clinical deployment may involve partial information. Finally, several variable-level comparisons had small denominators (e.g., hematocrit in PV clinical notes, n = 21), limiting the precision of those specific point estimates.

Future work will focus on prospective, multi-institution external validation across geographically and demographically diverse health systems; head-to-head evaluation of locally deployable and open-source LLMs against the GPT-4 Turbo reference to support broader institutional adoption;^32,33^ integration into operational clinical workflows for real-time decision support; and extension of the framework to other rare hematologic malignancies. Scalable phenotyping systems of this kind may also enable population-level research applications, including automated cohort assembly, identification of patients potentially eligible for clinical trials, and longitudinal monitoring of disease progression.^34,35^

## Conclusions

Automated phenotypic characterization of MPNs using an LLM-based framework is feasible and achieves expert-level performance for diagnostic classification and risk stratification across PV, ET, and MF. By automating a process that currently requires extensive manual review and subspecialty expertise, such systems may extend standardized care beyond specialized centers, enable scalable data infrastructure for research in rare hematologic malignancies, and provide a template that can be adapted to other rare hematologic malignancies with comparably complex, multi-source diagnostic and prognostic criteria. Prospective, multi-institution validation represents an important next step toward clinical implementation.

## Data Availability

De-identified analytic outputs can be made available to qualified investigators upon reasonable request and execution of an appropriate data use agreement, subject to institutional policy and HIPAA constraints. Source code for the analysis pipeline is available at https://github.com/muhammadali-k/mpn-phenotyping-pipeline.

## Acknowledgements

The authors thank the institutional research informatics team for support with secure access to the electronic health record data warehouse, and the clinical hematology service for contributions to ground-truth adjudication.

## Author contributions

All authors contributed to the study and approved the final manuscript.

