## Supplementary Material for "Automated Phenotypic Characterization in Rare Hematologic Malignancies Using a Large Language Model–Based Framework"

**Supplementary index**

Supplementary Table 1. Variable-level LLM extraction performance, by disease, document type, and variable.

Supplementary Table 2. Diagnostic classification performance compared with manual reference standard.

Supplementary Table 3. Prognostic risk-stratification performance compared with manual reference standard.

Supplementary Table 4. Source-ablation analysis: diagnostic and prognostic performance under varying source configurations.

Supplementary Figure 1. Source-prioritization decision tree for categorical variables (bone marrow features, karyotype, and mutation status).

Supplementary Figure 2. Source-prioritization decision tree for numeric laboratory variables.

Supplementary Figure 3. Polycythemia vera: concordance between structured laboratory values and pathology-report-extracted values.

Supplementary Figure 4. Essential thrombocythemia: concordance between structured laboratory values and pathology-report-extracted values.

Supplementary Figure 5. Myelofibrosis: concordance between structured laboratory values and pathology-report-extracted values.

Supplementary Figure 6. Source ablation: diagnostic sensitivity and specificity by source configuration.

Supplementary Figure 7. Source ablation: prognostic risk-stratification accuracy by source configuration.

### **Supplementary Table 1. Variable-level LLM extraction performance.**

| **Disease** | **Document type** | **Variable** | **Type** | **Observations** | **Accuracy, %** | **F1** | **Cohen’s κ** |
| --- | --- | --- | --- | --- | --- | --- | --- |
| PV | Pathology Reports | Erythropoietin | Numeric | 2 | 100.0 | 1.00 | 1.00 |
| PV | Pathology Reports | Hematocrit | Numeric | 6 | 100.0 | 1.00 | 1.00 |
| PV | Pathology Reports | Hemoglobin | Numeric | 132 | 100.0 | 1.00 | 1.00 |
| PV | Pathology Reports | Hypercellular marrow | Categorical | 142 | 99.3 | 0.99 | 0.98 |
| PV | Pathology Reports | JAK2 mutation | Categorical | 82 | 100.0 | 1.00 | 1.00 |
| PV | Pathology Reports | PV (hematologist-defined) | Categorical | 145 | 99.3 | 0.99 | 0.99 |
| PV | Pathology Reports | Percent cellularity | Numeric | 141 | 100.0 | 1.00 | 1.00 |
| PV | Pathology Reports | Trilineage hematopoiesis | Categorical | 144 | 96.5 | 0.98 | 0.78 |
| PV | Clinical Notes | Erythropoietin | Numeric | 7 | 100.0 | 1.00 | 1.00 |
| PV | Clinical Notes | Hematocrit | Numeric | 21 | 81.0 | 0.68 | 0.80 |
| PV | Clinical Notes | Hemoglobin | Numeric | 23 | 100.0 | 1.00 | 1.00 |
| PV | Clinical Notes | Hypercellular marrow | Categorical | 32 | 100.0 | 1.00 | 1.00 |
| PV | Clinical Notes | JAK2 mutation | Categorical | 39 | 100.0 | 1.00 | 1.00 |
| PV | Clinical Notes | PV (hematologist-defined) | Categorical | 52 | 94.2 | 0.96 | 0.86 |
| PV | Clinical Notes | Percent cellularity | Numeric | 21 | 95.2 | 0.90 | 0.95 |
| PV | Clinical Notes | Thrombotic history | Categorical | 52 | 98.1 | 0.96 | 0.95 |
| PV | Clinical Notes | Trilineage hematopoiesis | Categorical | 25 | 100.0 | 1.00 | 1.00 |
| ET | Pathology Reports | ASXL1 mutation | Categorical | 150 | 100.0 | 1.00 | 1.00 |
| ET | Pathology Reports | CALR mutation | Categorical | 150 | 100.0 | 1.00 | 1.00 |
| ET | Pathology Reports | ET (hematologist-defined) | Categorical | 150 | 100.0 | 1.00 | 1.00 |
| ET | Pathology Reports | EZH2 mutation | Categorical | 150 | 100.0 | 1.00 | 1.00 |
| ET | Pathology Reports | Hemoglobin | Numeric | 146 | 100.0 | 1.00 | 1.00 |
| ET | Pathology Reports | Hypercellular marrow | Categorical | 150 | 98.7 | 0.99 | 0.97 |
| ET | Pathology Reports | IDH1/2 mutation | Categorical | 150 | 100.0 | 1.00 | 1.00 |
| ET | Pathology Reports | JAK2 mutation | Categorical | 150 | 100.0 | 1.00 | 1.00 |
| ET | Pathology Reports | Lactate dehydrogenase | Numeric | 0 | — | — | — |
| ET | Pathology Reports | MPL mutation | Categorical | 150 | 100.0 | 1.00 | 1.00 |
| ET | Pathology Reports | Megakaryocyte atypia | Categorical | 150 | 98.0 | 0.98 | 0.96 |
| ET | Pathology Reports | Megakaryocyte proliferation | Categorical | 150 | 99.3 | 0.99 | 0.98 |
| ET | Pathology Reports | Percent cellularity | Numeric | 150 | 100.0 | 1.00 | 1.00 |
| ET | Pathology Reports | Platelet count | Numeric | 146 | 100.0 | 1.00 | 1.00 |
| ET | Pathology Reports | Reticulin fibrosis grade | Numeric | 93 | 98.9 | 0.99 | 0.98 |
| ET | Pathology Reports | SF3B1 mutation | Categorical | 150 | 99.3 | 0.80 | 0.80 |
| ET | Pathology Reports | SRSF2 mutation | Categorical | 150 | 100.0 | 1.00 | 1.00 |
| ET | Pathology Reports | TET2 mutation | Categorical | 150 | 100.0 | 1.00 | 1.00 |
| ET | Pathology Reports | White blood cell count | Numeric | 147 | 99.3 | 0.99 | 0.99 |
| ET | Clinical Notes | ASXL1 mutation | Categorical | 55 | 100.0 | 1.00 | 1.00 |
| ET | Clinical Notes | CALR mutation | Categorical | 55 | 100.0 | 1.00 | 1.00 |
| ET | Clinical Notes | ET (hematologist-defined) | Categorical | 55 | 100.0 | 1.00 | 1.00 |
| ET | Clinical Notes | EZH2 mutation | Categorical | 55 | 100.0 | 1.00 | 1.00 |
| ET | Clinical Notes | Hemoglobin | Numeric | 21 | 100.0 | 1.00 | 1.00 |
| ET | Clinical Notes | Hypercellular marrow | Categorical | 28 | 100.0 | 1.00 | 1.00 |
| ET | Clinical Notes | IDH1/2 mutation | Categorical | 55 | 100.0 | 1.00 | 1.00 |
| ET | Clinical Notes | JAK2 mutation | Categorical | 55 | 100.0 | 1.00 | 1.00 |
| ET | Clinical Notes | Lactate dehydrogenase | Numeric | 4 | 100.0 | 1.00 | 1.00 |
| ET | Clinical Notes | MPL mutation | Categorical | 55 | 100.0 | 1.00 | 1.00 |
| ET | Clinical Notes | Megakaryocyte atypia | Categorical | 19 | 100.0 | 1.00 | 1.00 |
| ET | Clinical Notes | Megakaryocyte proliferation | Categorical | 20 | 100.0 | 1.00 | 1.00 |
| ET | Clinical Notes | Percent cellularity | Numeric | 16 | 100.0 | 1.00 | 1.00 |
| ET | Clinical Notes | Platelet count | Numeric | 26 | 100.0 | 1.00 | 1.00 |
| ET | Clinical Notes | Reticulin fibrosis grade | Numeric | 22 | 100.0 | 1.00 | 1.00 |
| ET | Clinical Notes | SF3B1 mutation | Categorical | 55 | 100.0 | 1.00 | 1.00 |
| ET | Clinical Notes | SRSF2 mutation | Categorical | 55 | 100.0 | 1.00 | 1.00 |
| ET | Clinical Notes | Splenomegaly | Categorical | 38 | 100.0 | 1.00 | 1.00 |
| ET | Clinical Notes | TET2 mutation | Categorical | 55 | 100.0 | 1.00 | 1.00 |
| ET | Clinical Notes | Thrombotic history | Categorical | 49 | 100.0 | 1.00 | 1.00 |
| ET | Clinical Notes | White blood cell count | Numeric | 22 | 100.0 | 1.00 | 1.00 |
| MF | Pathology Reports | ASXL1 mutation | Categorical | 59 | 100.0 | 1.00 | 1.00 |
| MF | Pathology Reports | CALR Type 1 mutation | Categorical | 81 | 100.0 | 1.00 | 1.00 |
| MF | Pathology Reports | CALR mutation | Categorical | 85 | 100.0 | 1.00 | 1.00 |
| MF | Pathology Reports | EZH2 mutation | Categorical | 52 | 100.0 | 1.00 | 1.00 |
| MF | Pathology Reports | Hemoglobin | Numeric | 138 | 100.0 | 1.00 | 1.00 |
| MF | Pathology Reports | Hypercellular marrow | Categorical | 145 | 97.9 | 0.99 | 0.95 |
| MF | Pathology Reports | IDH1/2 mutation | Categorical | 58 | 100.0 | 1.00 | 1.00 |
| MF | Pathology Reports | JAK2 mutation | Categorical | 87 | 98.9 | 0.99 | 0.98 |
| MF | Pathology Reports | Lactate dehydrogenase | Numeric | 0 | — | — | — |
| MF | Pathology Reports | MF (hematologist-defined) | Categorical | 150 | 96.7 | 0.97 | 0.93 |
| MF | Pathology Reports | MPL mutation | Categorical | 76 | 100.0 | 1.00 | 1.00 |
| MF | Pathology Reports | Megakaryocyte atypia | Categorical | 146 | 100.0 | 1.00 | 1.00 |
| MF | Pathology Reports | Megakaryocyte proliferation | Categorical | 146 | 100.0 | 1.00 | 1.00 |
| MF | Pathology Reports | Percent cellularity | Numeric | 146 | 100.0 | 1.00 | 1.00 |
| MF | Pathology Reports | Peripheral blood blasts | Numeric | 147 | 100.0 | 1.00 | 1.00 |
| MF | Pathology Reports | Reactive fibrosis | Categorical | 92 | 98.9 | 0.80 | 0.79 |
| MF | Pathology Reports | Reticulin fibrosis grade | Numeric | 104 | 100.0 | 1.00 | 1.00 |
| MF | Pathology Reports | SF3B1 mutation | Categorical | 55 | 100.0 | 1.00 | 1.00 |
| MF | Pathology Reports | SRSF2 mutation | Categorical | 59 | 100.0 | 1.00 | 1.00 |
| MF | Pathology Reports | TET2 mutation | Categorical | 53 | 100.0 | 1.00 | 1.00 |
| MF | Pathology Reports | Unfavorable karyotype | Categorical | 111 | 99.1 | 0.98 | 0.97 |
| MF | Pathology Reports | Very-high-risk karyotype | Categorical | 110 | 100.0 | 1.00 | 1.00 |
| MF | Pathology Reports | White blood cell count | Numeric | 138 | 100.0 | 1.00 | 1.00 |
| MF | Clinical Notes | ASXL1 mutation | Categorical | 65 | 100.0 | 1.00 | 1.00 |
| MF | Clinical Notes | CALR Type 1 mutation | Categorical | 65 | 100.0 | 1.00 | 1.00 |
| MF | Clinical Notes | CALR mutation | Categorical | 65 | 100.0 | 1.00 | 1.00 |
| MF | Clinical Notes | Constitutional symptoms | Categorical | 61 | 100.0 | 1.00 | 1.00 |
| MF | Clinical Notes | EZH2 mutation | Categorical | 65 | 100.0 | 1.00 | 1.00 |
| MF | Clinical Notes | Hemoglobin | Numeric | 28 | 100.0 | 1.00 | 1.00 |
| MF | Clinical Notes | Hypercellular marrow | Categorical | 31 | 100.0 | 1.00 | 1.00 |
| MF | Clinical Notes | IDH1/2 mutation | Categorical | 65 | 100.0 | 1.00 | 1.00 |
| MF | Clinical Notes | JAK2 mutation | Categorical | 41 | 100.0 | 1.00 | 1.00 |
| MF | Clinical Notes | Lactate dehydrogenase | Numeric | 10 | 100.0 | 1.00 | 1.00 |
| MF | Clinical Notes | MF (hematologist-defined) | Categorical | 65 | 96.9 | 0.98 | 0.87 |
| MF | Clinical Notes | MPL mutation | Categorical | 65 | 100.0 | 1.00 | 1.00 |
| MF | Clinical Notes | Megakaryocyte atypia | Categorical | 24 | 100.0 | 1.00 | 1.00 |
| MF | Clinical Notes | Megakaryocyte proliferation | Categorical | 23 | 100.0 | 1.00 | 1.00 |
| MF | Clinical Notes | Percent cellularity | Numeric | 23 | 100.0 | 1.00 | 1.00 |
| MF | Clinical Notes | Peripheral blood blasts | Numeric | 35 | 100.0 | 1.00 | 1.00 |
| MF | Clinical Notes | Reactive fibrosis | Categorical | 13 | 92.3 | 0.67 | 0.63 |
| MF | Clinical Notes | Reticulin fibrosis grade | Numeric | 47 | 100.0 | 1.00 | 1.00 |
| MF | Clinical Notes | SF3B1 mutation | Categorical | 65 | 100.0 | 1.00 | 1.00 |
| MF | Clinical Notes | SRSF2 mutation | Categorical | 65 | 100.0 | 1.00 | 1.00 |
| MF | Clinical Notes | Splenomegaly | Categorical | 49 | 100.0 | 1.00 | 1.00 |
| MF | Clinical Notes | TET2 mutation | Categorical | 65 | 100.0 | 1.00 | 1.00 |
| MF | Clinical Notes | Unfavorable karyotype | Categorical | 40 | 100.0 | 1.00 | 1.00 |
| MF | Clinical Notes | Very-high-risk karyotype | Categorical | 39 | 100.0 | 1.00 | 1.00 |
| MF | Clinical Notes | White blood cell count | Numeric | 30 | 100.0 | 1.00 | 1.00 |

***Supplementary Table 1:*** *Per-variable extraction performance for the LLM-based pipeline against the manually adjudicated reference standard, stratified by disease, document type (pathology reports vs. clinical notes), and variable. Categorical variables were evaluated by exact-match agreement; numeric variables were evaluated as text-extraction tasks (exact-string match after canonical formatting). Per-variable values are reported as point estimates with paired-observation denominators (Observations); accuracy is reported as a percentage (0–100%) and F1 score is reported on the conventional 0–1.00 scale. 95% confidence intervals are reported only for aggregate disease- and patient-level metrics (Figure 2 and Supplementary Tables 2–4). By convention, F1 is set to 1.00 for fully concordant or all-negative configurations. Cells marked with “—” correspond to variables that were not reported in the corresponding document type during the available retrieval window. Several variables — most prominently serum erythropoietin and lactate dehydrogenase — appear in only a small number of pathology reports because they are serum analytes rather than bone-marrow findings and are infrequently re-transcribed into the morphology narrative; in these rows, the small number of observations should be interpreted as a feature of clinical documentation rather than a limitation of the extraction pipeline.* ***Abbreviations:*** *ET, essential thrombocythemia; F1, F1 score; LLM, large language model; MF, myelofibrosis; PV, polycythemia vera; κ, Cohen’s kappa.*

### **Supplementary Table 2. Diagnostic performance versus manual reference standard.**

| **Disease** | **N** | **Sensitivity, % (95% CI)** | **Specificity, % (95% CI)** | **Cohen’s κ** |
| --- | --- | --- | --- | --- |
| Polycythemia vera | 150 | 100.0 (95.1–100.0) | 98.7 (92.8–99.8) | 0.99 |
| Essential thrombocythemia | 150 | 100.0 (95.1–100.0) | 100.0 (95.1–100.0) | 1.00 |
| Myelofibrosis | 150 | 100.0 (95.1–100.0) | 100.0 (95.1–100.0) | 1.00 |

***Supplementary Table 2:*** *Patient-level diagnostic performance for the LLM-based pipeline against the manually adjudicated reference standard in the held-out test set (n = 150 per disease; 75 cases and 75 controls). Sensitivity, specificity, and Cohen’s kappa are reported as the primary diagnostic metrics; positive and negative predictive values, accuracy, and F1 score are not reported because the case-to-control sampling ratio was fixed at 1:1, which does not reflect the population prevalence of MPN and therefore renders prevalence-dependent metrics non-informative for clinical practice. 95% confidence intervals are Wilson intervals.* ***Abbreviations:*** *95% CI, 95% confidence interval; LLM, large language model; κ, Cohen’s kappa.*

### **Supplementary Table 3. Prognostic risk-stratification performance versus manual reference standard.**

| **Disease** | **N cases** | **Accuracy, % (95% CI)** | **Weighted F1 (95% CI)** | **Quadratic-weighted κ** | **Unassigned by LLM (n)** |
| --- | --- | --- | --- | --- | --- |
| Polycythemia vera | 75 | 100.0 (95.1–100.0) | 1.00 (1.00–1.00) | 1.00 | 0 |
| Essential thrombocythemia | 75 | 100.0 (95.1–100.0) | 1.00 (1.00–1.00) | 1.00 | 0 |
| Myelofibrosis | 75 | 100.0 (95.1–100.0) | 1.00 (1.00–1.00) | 1.00 | 0 |

***Supplementary Table 3:*** *Prognostic risk-stratification performance among confirmed cases (n = 75 per disease) against the manually adjudicated reference standard. PV was stratified using the conventional risk model; ET was stratified using the four-tier IPSET-thrombosis system; MF was stratified using DIPSS, DIPSS-plus, and MIPSS70/MIPSS70+ v2, with model selection driven by the availability of cytogenetic and high-molecular-risk mutation data. Accuracy is reported as a percentage (0–100%) and weighted F1 score is reported on the conventional 0–1.00 scale. Quadratic-weighted κ was used to reflect the ordinal nature of the risk categories. 95% confidence intervals are Wilson intervals (proportions) or bootstrap intervals from 500 resamples (F1).* ***Abbreviations:*** *95% CI, 95% confidence interval; DIPSS, Dynamic International Prognostic Scoring System; ET, essential thrombocythemia; F1, F1 score; IPSET, International Prognostic Score for Thrombosis in Essential Thrombocythemia; LLM, large language model; MF, myelofibrosis; MIPSS70, Mutation-Enhanced International Prognostic Score System; PV, polycythemia vera; κ, Cohen’s kappa.*

### **Supplementary Table 4. Source-ablation analysis.**

| **Disease** | **Source configuration** | **N (diagnosis)** | **Sensitivity, % (95% CI)** | **Specificity, % (95% CI)** | **N (prognosis)** | **Prognostic accuracy, % (95% CI)** |
| --- | --- | --- | --- | --- | --- | --- |
| PV | Pathology only | 150 | 98.7 (92.8–99.8) | 100.0 (95.1–100.0) | 75 | 69.3 (58.2–78.6) |
| PV | Pathology + clinical notes | 150 | 98.7 (92.8–99.8) | 98.7 (92.8–99.8) | 75 | 100.0 (95.1–100.0) |
| PV | Pathology + structured labs | 150 | 100.0 (95.1–100.0) | 100.0 (95.1–100.0) | 75 | 69.3 (58.2–78.6) |
| PV | All sources | 150 | 100.0 (95.1–100.0) | 98.7 (92.8–99.8) | 75 | 100.0 (95.1–100.0) |
| ET | Pathology only | 150 | 100.0 (95.1–100.0) | 100.0 (95.1–100.0) | 75 | 93.3 (85.3–97.1) |
| ET | Pathology + clinical notes | 150 | 100.0 (95.1–100.0) | 100.0 (95.1–100.0) | 75 | 100.0 (95.1–100.0) |
| ET | Pathology + structured labs | 150 | 100.0 (95.1–100.0) | 100.0 (95.1–100.0) | 75 | 93.3 (85.3–97.1) |
| ET | All sources | 150 | 100.0 (95.1–100.0) | 100.0 (95.1–100.0) | 75 | 100.0 (95.1–100.0) |
| MF | Pathology only | 150 | 96.0 (88.9–98.6) | 100.0 (95.1–100.0) | 75 | 77.3 (66.7–85.3) |
| MF | Pathology + clinical notes | 150 | 96.0 (88.9–98.6) | 100.0 (95.1–100.0) | 75 | 100.0 (95.1–100.0) |
| MF | Pathology + structured labs | 150 | 100.0 (95.1–100.0) | 100.0 (95.1–100.0) | 75 | 82.7 (72.6–89.6) |
| MF | All sources | 150 | 100.0 (95.1–100.0) | 100.0 (95.1–100.0) | 75 | 100.0 (95.1–100.0) |

***Supplementary Table 4:*** *Source-ablation analysis evaluating diagnostic and prognostic performance under four progressively richer source configurations: pathology only; pathology + clinical notes; pathology + structured labs; and all sources combined. Diagnostic performance was largely insensitive to source configuration (sensitivity ≥ 96.0% across all configurations). Prognostic performance, in contrast, was substantially impaired without clinical notes: pathology-only and pathology + structured-labs configurations yielded prognostic accuracy of 69%–93%, whereas adding clinical notes restored accuracy to 100% across all three diseases. 95% confidence intervals are Wilson intervals.* ***Abbreviations:*** *95% CI, 95% confidence interval; ET, essential thrombocythemia; LLM, large language model; MF, myelofibrosis; PV, polycythemia vera.*

### **Supplementary Figure 1. Source-prioritization decision tree for categorical variables.**

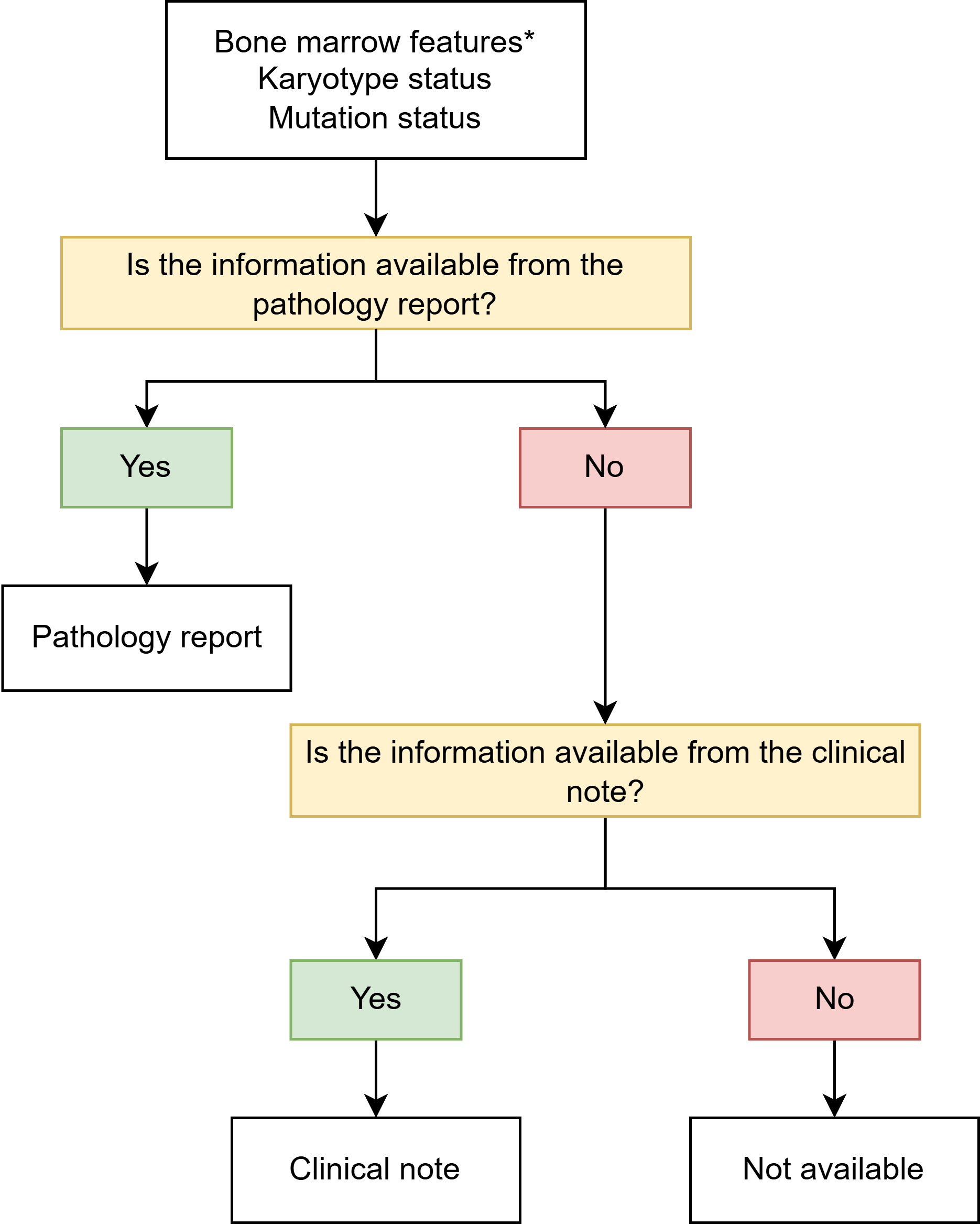

***Supplementary Figure 1:*** *Decision tree implementing the source-prioritization logic for categorical variables, including bone marrow morphologic features (hypercellularity, megakaryocyte proliferation and atypia, reticulin fibrosis grade, and trilineage hematopoiesis), karyotype status, and mutation status. The pathology report is queried first; if the requested information is not available there, the corresponding clinical note is queried; if not available in either source, the variable is recorded as “Not available.” *Bone marrow features comprise the morphologic variables listed above and are reported predominantly in pathology reports.*

### **Supplementary Figure 2. Source-prioritization decision tree for numeric laboratory variables.**

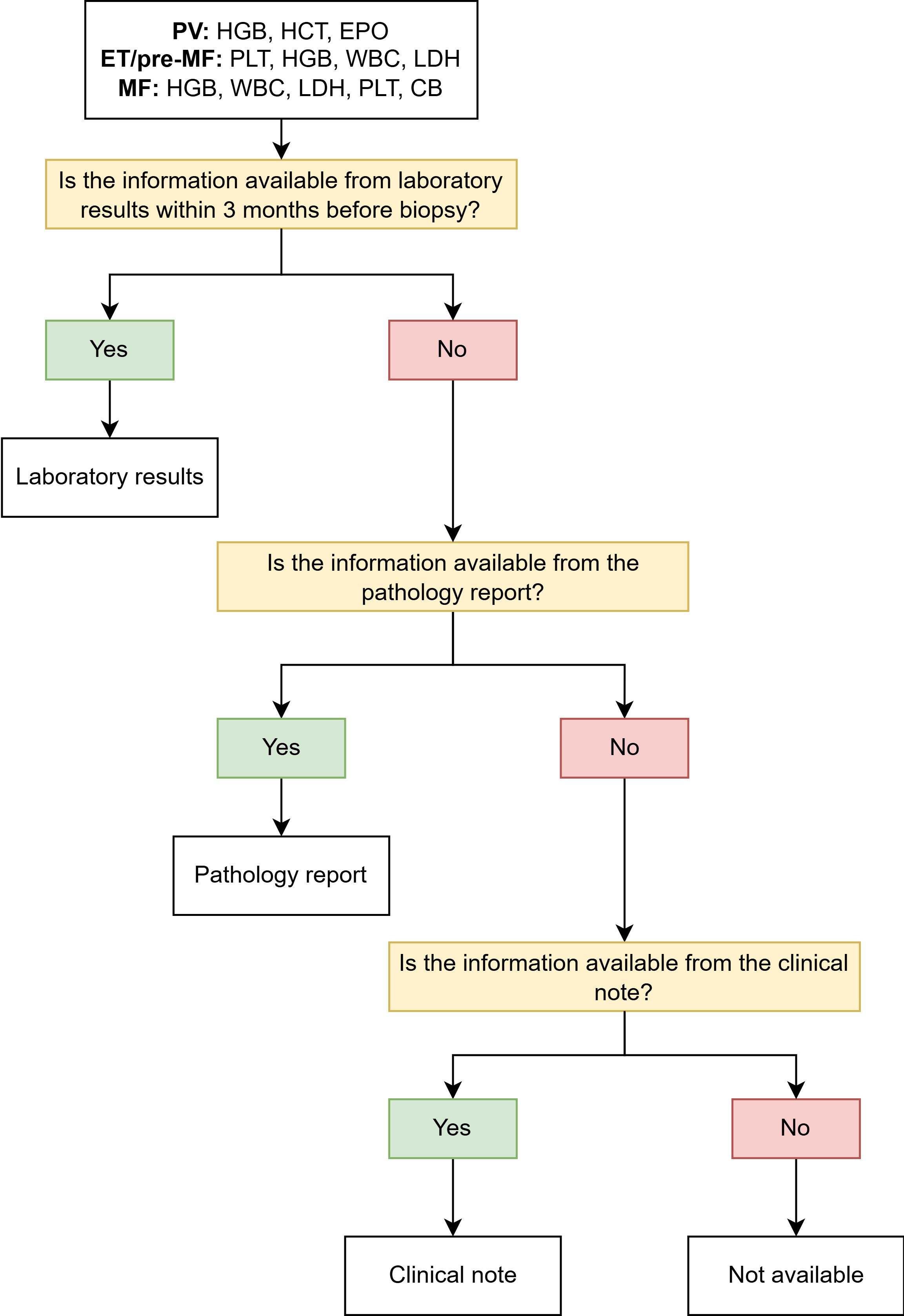

***Supplementary Figure 2:*** *Decision tree implementing the source-prioritization logic for numeric laboratory variables required by the diagnostic and prognostic models. For PV, hemoglobin (HGB), hematocrit (HCT), and serum erythropoietin (EPO) are sought; for ET and pre-fibrotic MF, platelet count (PLT), HGB, white blood cell count (WBC), and lactate dehydrogenase (LDH); for overt MF, HGB, WBC, LDH, PLT, and circulating peripheral blood blasts (CB). For each variable the structured laboratory record within 3 months prior to bone marrow biopsy is queried first; if not available, the pathology report is queried; if still not available, the corresponding clinical note is queried; if not available in any source, the variable is recorded as “Not available.”* ***Abbreviations:*** *CB, circulating peripheral blood blasts; EPO, erythropoietin; ET, essential thrombocythemia; HCT, hematocrit; HGB, hemoglobin; LDH, lactate dehydrogenase; MF, myelofibrosis; PLT, platelet count; PV, polycythemia vera; WBC, white blood cell count.*

### **Supplementary Figure 3. Polycythemia vera: concordance between structured laboratory values and pathology-report-extracted values.**

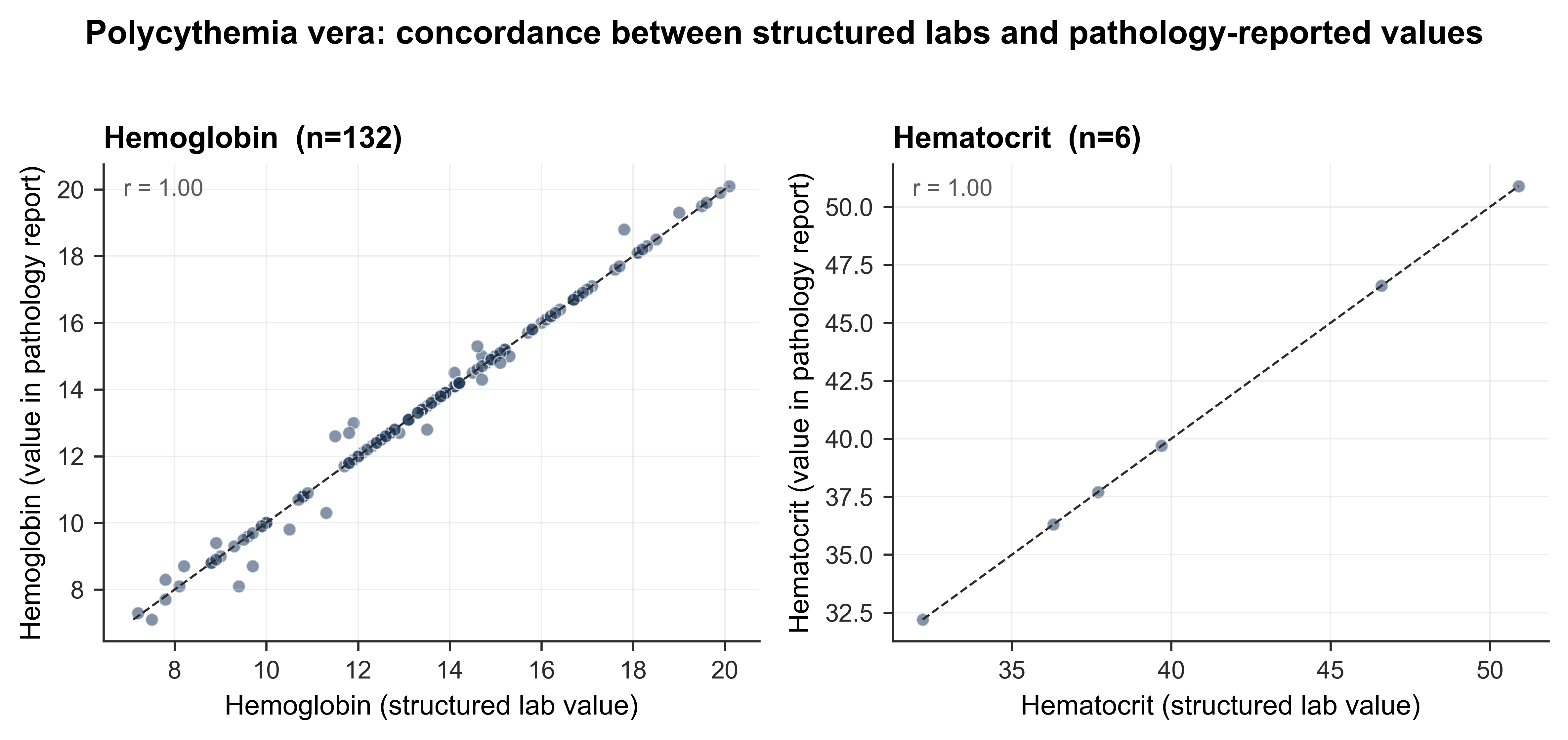

***Supplementary Figure 3:*** *Scatterplots of structured laboratory values (x-axis) versus the corresponding LLM-extracted values from the pathology report (y-axis) for hemoglobin (left) and hematocrit (right) in polycythemia vera. Dashed line is the line of identity; r is the Pearson correlation coefficient. Pearson r = 1.00 for both analytes.* ***Abbreviation:*** *LLM, large language model.*

### **Supplementary Figure 4. Essential thrombocythemia: concordance between structured laboratory values and pathology-report-extracted values.**

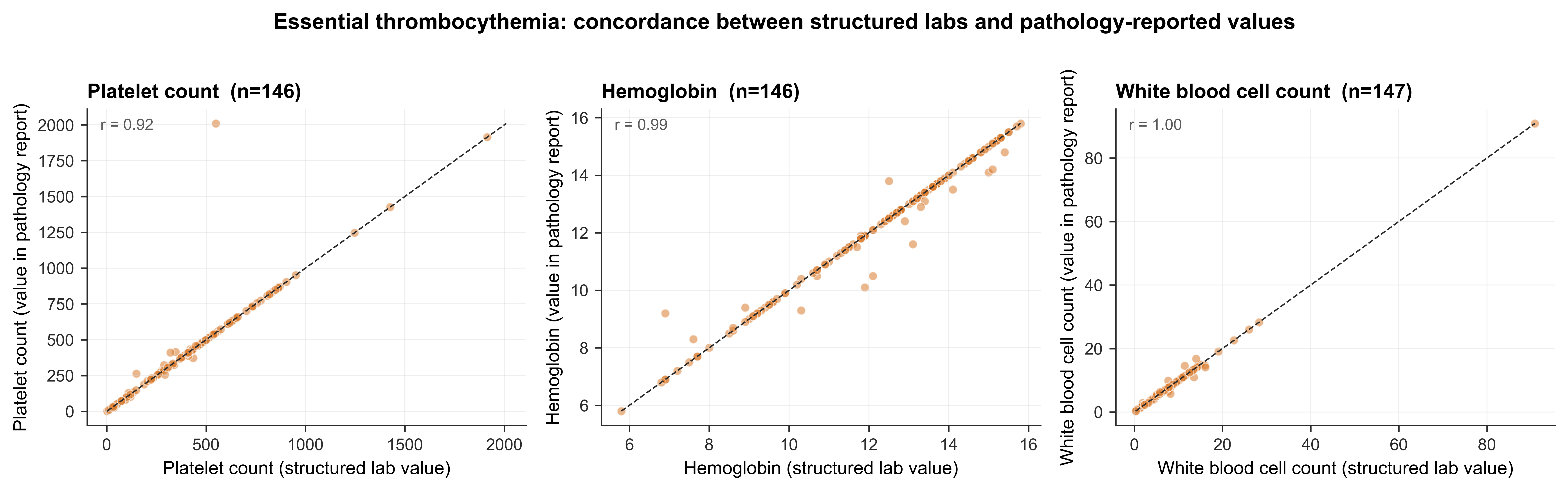

***Supplementary Figure 4:*** *Scatterplots of structured laboratory values (x-axis) versus LLM-extracted values from the pathology report (y-axis) for platelet count (left), hemoglobin (middle), and white blood cell count (right) in essential thrombocythemia. Dashed line is the line of identity; r is the Pearson correlation coefficient. Pearson r = 0.92 for platelet count, 0.99 for hemoglobin, and 1.00 for white blood cell count.* ***Abbreviation:*** *LLM, large language model.*

### **Supplementary Figure 5. Myelofibrosis: concordance between structured laboratory values and pathology-report-extracted values.**

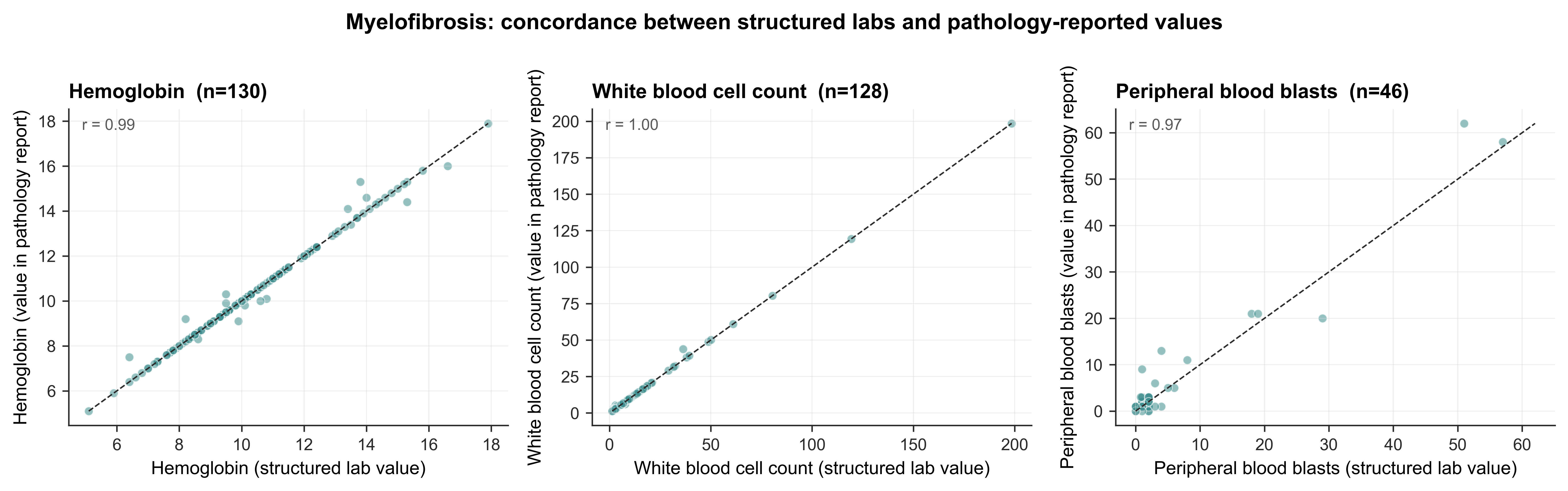

***Supplementary Figure 5:*** *Scatterplots of structured laboratory values (x-axis) versus LLM-extracted values from the pathology report (y-axis) for hemoglobin (left), white blood cell count (middle), and peripheral blood blasts (right) in myelofibrosis. Dashed line is the line of identity; r is the Pearson correlation coefficient. Pearson r = 0.99 for hemoglobin, 1.00 for white blood cell count, and 0.97 for peripheral blood blasts.* ***Abbreviation:*** *LLM, large language model.*

### **Supplementary Figure 6. Source ablation: diagnostic sensitivity and specificity.**

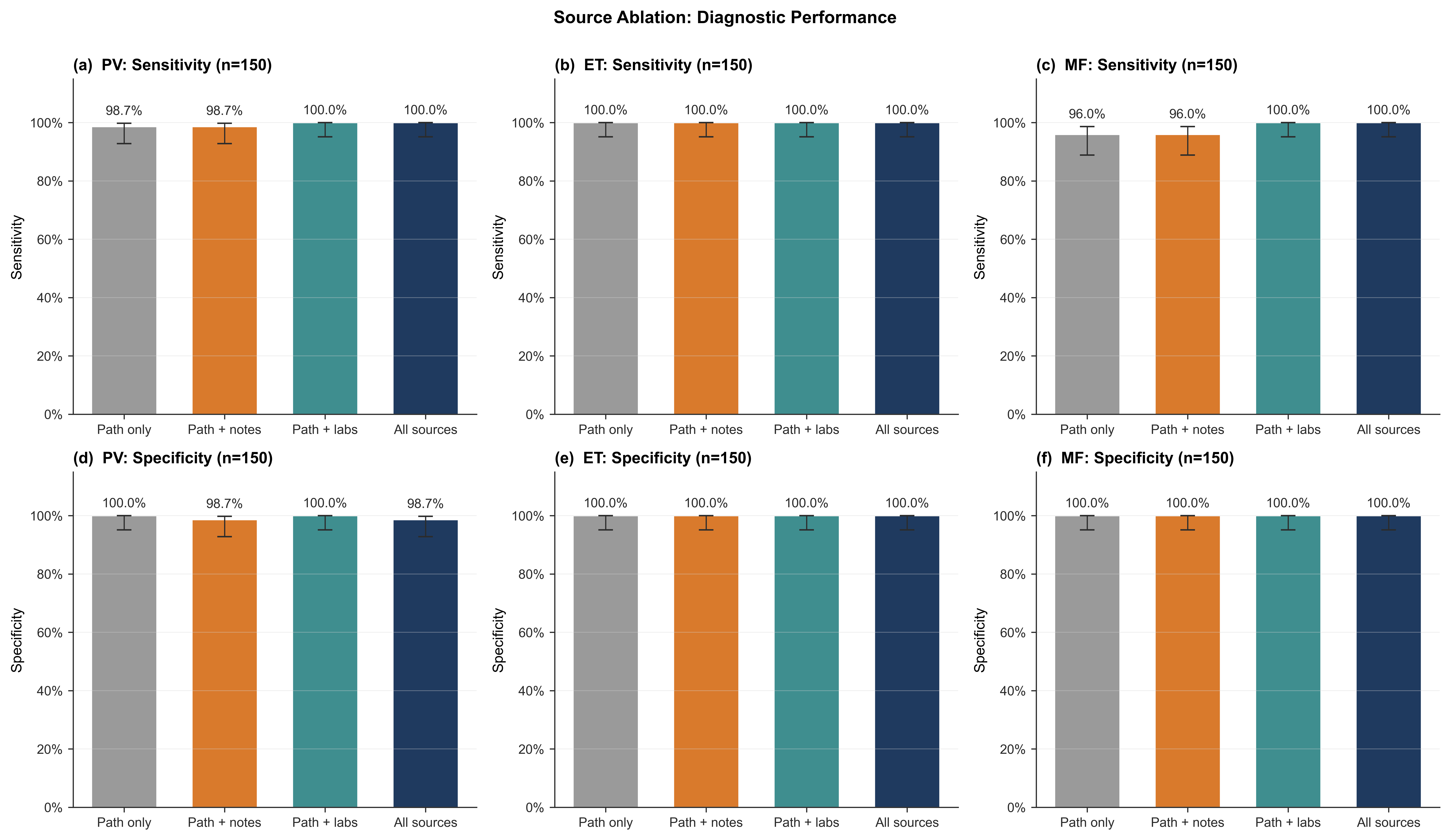

***Supplementary Figure 6:*** *Diagnostic sensitivity (top row, panels a–c) and specificity (bottom row, panels d–f) under four source configurations: pathology only (gray), pathology + clinical notes (orange), pathology + structured labs (teal), and all sources (navy), shown for PV (a, d), ET (b, e), and MF (c, f). Numbers above each bar give the point estimate; error bars indicate the 95% Wilson confidence interval.* ***Abbreviations:*** *ET, essential thrombocythemia; LLM, large language model; MF, myelofibrosis; PV, polycythemia vera.*

### **Supplementary Figure 7. Source ablation: prognostic risk-stratification accuracy.**

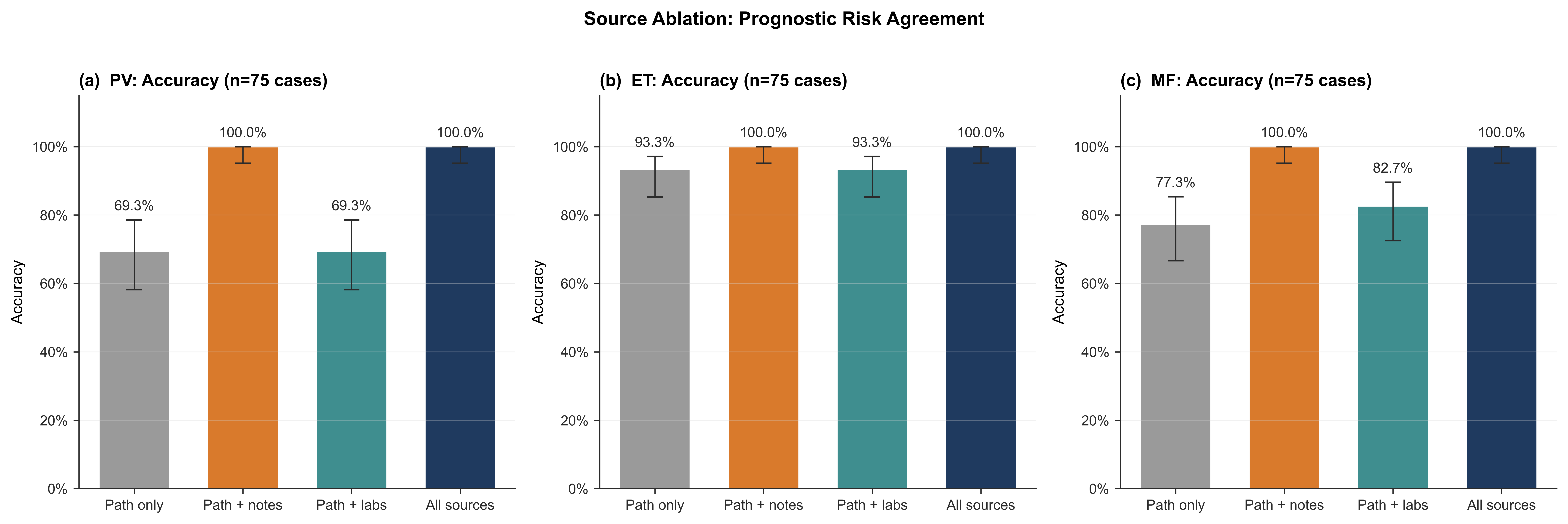

***Supplementary Figure 7:*** *Prognostic risk-stratification accuracy across four source configurations among confirmed cases (n = 75 per disease) for PV (a), ET (b), and MF (c). Bars show point estimates with Wilson 95% confidence intervals. Adding clinical notes to pathology reports restored accuracy to 100% across all three diseases; adding structured laboratory data alone did not.* ***Abbreviations:*** *ET, essential thrombocythemia; LLM, large language model; MF, myelofibrosis; PV, polycythemia vera.*
